# Identification and management of thyroid dysfunction using at-home sample collection and telehealth services

**DOI:** 10.1101/2022.09.29.22280520

**Authors:** Kathleen M. Gavin, Daniel Kreitzberg, Yvette Gaudreau, Marisa Cruz, Timothy A. Bauer

## Abstract

**Introduction:** Programs aimed at modernizing thyroid care by pairing at-home sample collection methods with telehealth options may serve an important and emerging role in thyroid care. The primary objective of this analysis was to evaluate telehealth utilization, demographics, and clinical characteristics of a cohort of consumer initiated at-home lab thyroid test users who were also offered the option of follow-up telehealth consultations.

**Methods:** This was a retrospective analysis of real-world data from a de-identified consumer database of home-collected, mail-in Thyroid Tests utilized from March to May 2021 (n=8,152). The mean age was 38.6 (range 18-85) years and 86.6% of individuals identified as female.

**Results:** Seven percent of test takers fell into a thyroid dysfunction category (0.9% overt-hypothyroidism, 2.9% subclinical hypothyroidism, 0.1% overt-hypothyroidism, and 3.3% subclinical-hyperthyroidism). Twelve percent of the overall sample opted into a telehealth consultation, with 91.8% receiving a non-treatment telehealth consultation and 8.2% receiving a treatment telemedicine consultation. Sixteen percent of individuals with overt or subclinical thyroid dysfunction engaged in telehealth consultations. Of those opting into a treatment consultation, 59.3% reported a history of thyroid issues, 55.6% indicated wanting to discuss their current thyroid medication, and 48% received a prescription medication.

**Discussion:** The combination of at-home sample collection and telehealth is an innovative model for screening for thyroid disorders, monitoring thyroid function, and increasing access to care that can be implemented at a large scale and across a wide range of age groups.

## Introduction

Telehealth is an attractive option for modern healthcare as it has the potential to improve many barriers to care including accessibility, quality, and cost.^1^ Research suggests patients with a wide variety of health problems, including thyroid disorders, are generally satisfied with health management via telemedicine.^2-4^ Although to date it’s use has been limited in scope, telemedicine has the potential to serve an important and emerging role for managing thyroid disorders,^5, 6^ particularly in combination with laboratory testing. Detection and diagnosis of thyroid disorders is an important public health issue. According to the American Thyroid Association, nearly 20 million Americans currently have some form of thyroid disease and over 12% of individuals in the United States (U.S.) will develop a thyroid condition within their lifetime.^7^

It is common for patients to request specific tests from their physicians.^8-10^ More recently, commercial avenues have become available for consumers to pursue testing. In consumer-initiated testing (CIT), individuals order a lab test based on their health-related needs and/or goals, which is then reviewed and authorized by a physician. In some cases, the associated sample collection can be completed by the consumer at home. Combining at-home sample collection with certified lab-based testing and telehealth services can provide an accessible and convenient platform for consumers to initiate management of their health. In resource-limited settings, or during periods in which travel is restricted or healthcare resources overwhelmed, these approaches may be particularly appropriate.^11^

Studies suggest that those with symptoms of thyroid dysfunction may have lower quality of life and higher rates of clinical comorbidities such as cardiovascular diseases and diabetes.^12, 13^ Symptoms of hypo-or hyperthyroidism (e.g., fatigue, anxiety, depression, impaired memory, unintentional weight loss and/or irregular menstrual periods) may lead an individual to pursue CIT for thyroid health evaluation. However, limited data are available regarding novel programs aimed at modernizing thyroid care utilizing a combination of CIT with at-home collection methods and telehealth offerings. The primary objective of this analysis was to evaluate telehealth utilization, demographics, and clinical characteristics of a cohort in a pilot program pairing consumer-initiated home-collection and lab-based testing with the option for follow-up telehealth consultations.

## Methods

This is a retrospective data analysis of real-world data. The project was deemed exempt from IRB review by WCG IRB because it does not meet the definition of human subjects research as defined in federal regulation 45 CFR 46.102.

### Data collection and testing

This analysis included de-identified data from a consumer database of at-home collected, mail-in Thyroid Tests (Everlywell, Inc., Austin, TX). Consumers had independently purchased thyroid test collection kits. Data from consumers aged 18 and older and with valid test results collected between March 1, 2021, and May 12, 2021, were eligible for inclusion in the analysis. Age, sex, and zip code were reported at purchase. Truncated zip codes were used to categorize U.S. census regions.

The Thyroid test collection kits included all materials necessary and instructions for self-collection of dry blood spot (DBS) samples, including a link to an instructional video on how to properly perform the finger stick, the self-collection procedure, and how to evaluate the sample sufficiency. Following collection, DBS samples were shipped, using prepaid shipping materials, to CLIA/CAP-certified Everlywell Diagnostics (EWDx, Dallas, TX) or CLIA-certified ZRT Laboratory (ZRT, Beaverton, OR) for analysis of Thyroid-Stimulating Hormone (TSH), free thyroxine (T_4_), free Triiodothyronine (T_3_), and Thyroid Peroxidase Antibodies (TPO). Only TSH and free T_4_ were included in this analysis. The lab-defined normal range for TSH was 0.55 to 4.78 mIU/mL at EWDx and 0.5 to 3.0 mIU/mL at ZRT. The normal range for free T_4_ was 0.7 to 2.5 ng/dL at both laboratories. Kits processed at ZRT with TSH results below 0.2 mIU/mL were reported as “<0.2” (n=14) and included in the analysis as 0.1 mIU/mL. Test results otherwise outside of the limits of detection and tests that did not result were excluded from the analysis.

Notification emails were sent to patients when their results were available. A digital platform displayed biomarker results in the context of the lab-defined normal ranges and included an opt-in link for a telehealth follow-up with a healthcare professional for interested individuals. The telehealth intake form included an eligibility questionnaire. If eligibility requirements for a treatment consultation were not met, patients were given the option to continue with a non-treatment telehealth consultation. Patients with a TSH result > 10 mIU/mL, were contacted via telephone by a patient care department representative. During the phone call, patients were notified that they received an abnormal TSH result and that a physician was available to answer questions and help manage their thyroid health. The average time from sample collection to telehealth consultation was 11.1 days (s.d.=4.3, range 3-27 days). Time from results approved and available to patient to consultation was on average 2.0 days (s.d.=3.5, range 0-14). Twenty-three different physicians provided both treatment and non-treatment consultations including a median of 84 consultations per physician, ranging from 1 to 227 consultations per physician. Physician specialties included 52.2% family medicine, 30.4% internal medicine, 13.0% emergency medicine and/or preventative medicine.

Eligibility for telemedicine treatment consultations included having a TSH value ≥ 4 mIU/mL or currently taking any thyroid medication. Exclusion criteria included: pregnant, planning to become pregnant, or within 1-year postpartum; Turner syndrome; Down syndrome; multiple sclerosis; ischemic heart disease; goiter or suspected goiter; disorders affecting pituitary gland or hypothalamus; endocrine dysfunction as a result of traumatic brain injury; ongoing cancer treatment; HIV; received contrast in the past 6 weeks; or currently taking lithium carbonate, amiodarone, aminoglutethimide, interferon L, thalidomide, betaroxine, or stavudine.

### Statistical methods

Descriptive statistics were used to summarize the study sample including demographics, prevalence of individuals within thyroid function categories, range of biomarker values within thyroid function categories, the proportions of individuals who opted for treatment or non-treatment consultations, and prescription status. Descriptive statistics include number and percentage for categorical variables and medians and interquartile ranges or means and standard deviations for continuous variables. All statistics were calculated in R (version 4.0.5) for Macintosh.

## Results

The analysis included test results from 8,152 individuals (Table 1). Over half of the sample was 30 to 44 years old, with nearly 90% identifying as female. Seven percent of test takers fell into a thyroid dysfunction category and the remaining 93% of the sample was categorized as euthyroid.

**Table 1.** Demographic characteristics.

| **Participant Characteristics** | **Total N (%)**<br>**(N = 8,152, 100%)** |
| --- | --- |
| **Age** |  |
| 18 - 29 years | 1,661 (20.4%) |
| 30 - 44 years | 4,449 (54.6%) |
| 45 - 55 years | 1,363 (16.7%) |
| 56 - 69 years | 585 (7.2%) |
| ≥ 70 years | 94 (1.2%) |
| **Sex** |  |
| Male | 1,091 (13.4%) |
| Female | 7,061 (86.6%) |

Data are the number and percentage of all Thyroid test users.

The TSH and free T_4_ values among thyroid function categories are in Table 2. As expected, the range of TSH and free T_4_ values correspond with criteria for identifying each thyroid function category. Individuals in the euthyroid category had a median (IQR) TSH level of 1.45 (0.9) mIU/L and mean (s.d.) free T_4_ value of 12.5 (3.7) pmol/L. In general, the difference in proportion between males and females within each thyroid function category tracked with overall test usage, with females making up ≥ 83% of those in each category.

**Table 2.**
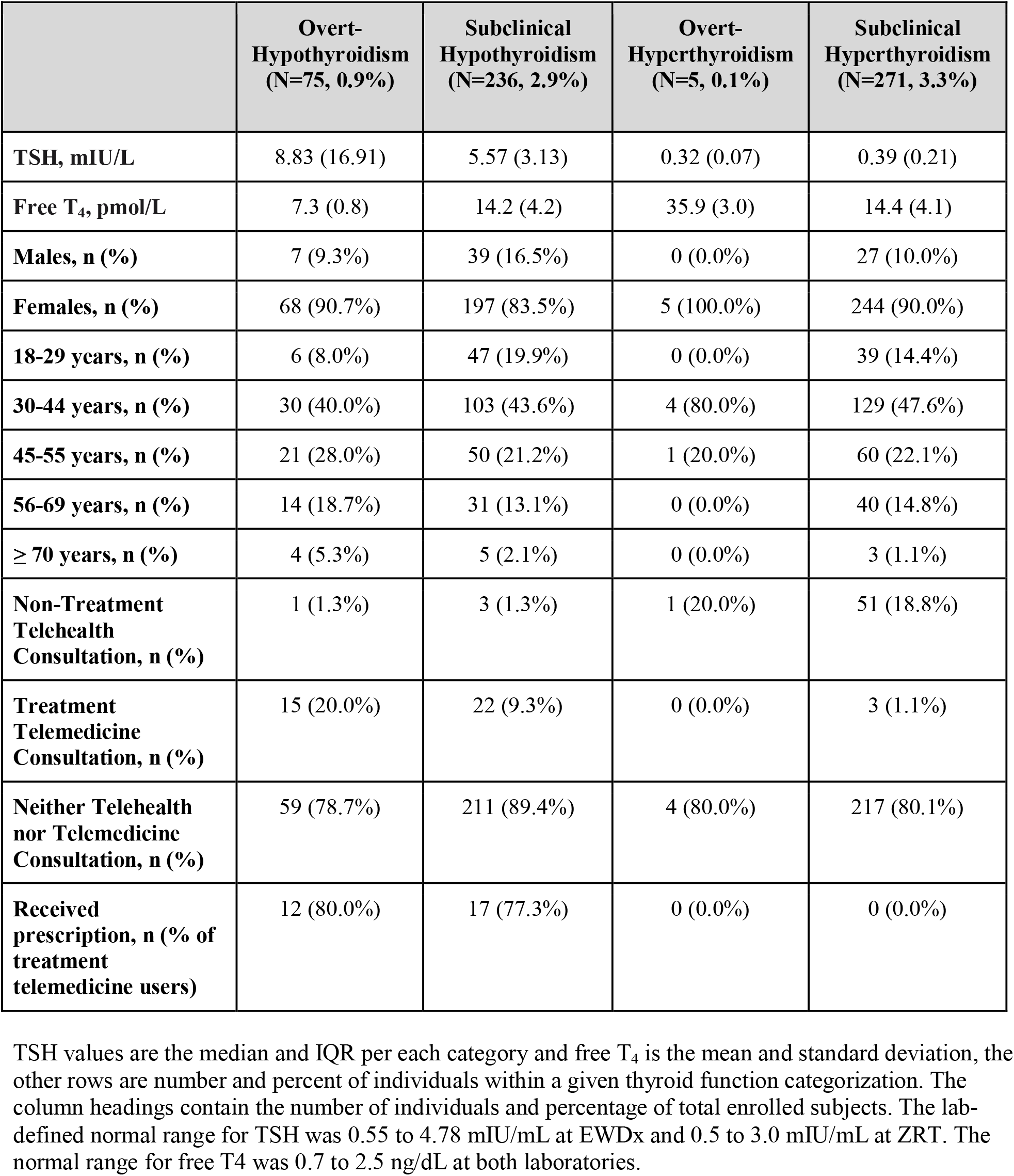
Clinical thyroid categories and telemedicine opt-ins.

|  | **Overt-Hypothyroidism**<br>**(N=75, 0.9%)** | **Subclinical Hypothyroidism**<br>**(N=236, 2.9%)** | **Overt-Hyperthyroidism**<br>**(N=5, 0.1%)** | **Subclinical Hyperthyroidism**<br>**(N=271, 3.3%)** |
| --- | --- | --- | --- | --- |
| **TSH, mIU/L** | 8.83 (16.91) | 5.57 (3.13) | 0.32 (0.07) | 0.39 (0.21) |
| **Free T<sub>4</sub>, pmol/L** | 7.3 (0.8) | 14.2 (4.2) | 35.9 (3.0) | 14.4 (4.1) |
| **Males, n (%)** | 7 (9.3%) | 39 (16.5%) | 0 (0.0%) | 27 (10.0%) |
| **Females, n (%)** | 68 (90.7%) | 197 (83.5%) | 5 (100.0%) | 244 (90.0%) |
| **18-29 years, n (%)** | 6 (8.0%) | 47 (19.9%) | 0 (0.0%) | 39 (14.4%) |
| **30-44 years, n (%)** | 30 (40.0%) | 103 (43.6%) | 4 (80.0%) | 129 (47.6%) |
| **45-55 years, n (%)** | 21 (28.0%) | 50 (21.2%) | 1 (20.0%) | 60 (22.1%) |
| **56-69 years, n (%)** | 14 (18.7%) | 31 (13.1%) | 0 (0.0%) | 40 (14.8%) |
| **≥ 70 years, n (%)** | 4 (5.3%) | 5 (2.1%) | 0 (0.0%) | 3 (1.1%) |
| **Non-Treatment Telehealth Consultation, n (%)** | 1 (1.3%) | 3 (1.3%) | 1 (20.0%) | 51 (18.8%) |
| **Treatment Telemedicine Consultation, n (%)** | 15 (20.0%) | 22 (9.3%) | 0 (0.0%) | 3 (1.1%) |
| **Neither Telehealth nor Telemedicine Consultation, n (%)** | 59 (78.7%) | 211 (89.4%) | 4 (80.0%) | 217 (80.1%) |
| **Received prescription, n (% of treatment telemedicine users)** | 12 (80.0%) | 17 (77.3%) | 0 (0.0%) | 0 (0.0%) |

TSH values are the median and IQR per each category and free T<sub>4</sub> is the mean and standard deviation, the other rows are number and percent of individuals within a given thyroid function categorization. The column headings contain the number of individuals and percentage of total enrolled subjects. The lab-defined normal range for TSH was 0.55 to 4.78 mIU/mL at EWDx and 0.5 to 3.0 mIU/mL at ZRT. The normal range for free T<sub>4</sub> was 0.7 to 2.5 ng/dL at both laboratories.

Hypothyroid categorization aligned with increased prevalence in older age, with those ≥ 45 years of age making up 52% of individuals categorized as overt-hypothyroid and 36% of those categorized as subclinical hypothyroid (Table 2). However, as would be expected, 80% of individuals in the overt-hyperthyroidism category and 62% in the subclinical hyperthyroid categories were < 45 years of age.

Twelve percent of the overall sample opted into any telehealth consultation (range by thyroid function categorization, 10.6%-21.3%, Table 2). Among patients who received a TSH result ≥ 4 mIU/L (n=376), 13.8% (n=52) opted-in for a telehealth consultation and 88.5% (n=46) of those were treatment consultations. In addition, of those with overt or subclinical thyroid dysfunction (n=587), 16.4% (n=96) engaged in telehealth consultations, 6.8% (n=40) of which were treatment consultations. The non-treatment telehealth consultation was primarily utilized by individuals in the subclinical and overt-hyperthyroid (Table 2) and euthyroid (n=847, 11.2% of category) categories who did not qualify for a treatment consultation.

Table 3 describes the telehealth users by consultation type. Twenty-one percent of those engaging in any telehealth consultation had a pre-existing thyroid condition, while the other 79% were de novo. The most widely reported reason for opting into a telehealth consultation overall was to discuss test results (88.9%), followed by wanting to understand how to treat thyroid dysfunction (53.5%). While individuals with a history of thyroid issues only represented 17.4% of non-treatment telehealth consultations, they represented over half of the treatment telemedicine group. There was greater representation from older age groups opting into treatment consultations compared to the overall sample, which was reflective of their greater proportion in the hypothyroid categories. Medication was of importance to those in the treatment telemedicine group; 55.6% indicated being interested in discussing their current medication and 48.1% (n=39) received a prescription medication, 43.6% of whom indicated starting a medication as the reason for wanting a consultation. Notably, half of those who received a prescription medication reported a history of thyroid issues. Of the 83 individuals who were currently taking a thyroid medication and wanted to discuss potential changes, 31.3% had biomarker levels indicative of thyroid dysfunction, 54.2% received a telemedicine treatment consultation, and 15.6% received a prescription medication from that consultation.

**Table 3.** Characterization of individuals completing either a non-treatment telehealth or treatment telemedicine consultation.

|  | **Non-Treatment Consultation\***<br>(N=903, 91.8%) | **Treatment Consultation†**<br>(N=81, 8.2%) | **Received Rx‡**<br>(N=39, 4.0%) | **Total**<br>(N=984, 100.0%) |
| --- | --- | --- | --- | --- |
| **Demographics** |  |  |  |  |
| Female | 786 (87.0%) | 79 (97.5%) | 38 (97.4%) | 865 (87.9%) |
| **Age** |  |  |  |  |
| 18-29 years | 212 (23.5%) | 11 (13.6%) | 7 (17.9%) | 223 (22.7%) |
| 30-44 years | 510 (56.5%) | 32 (39.5%) | 15 (38.5%) | 542 (55.1%) |
| 45 - 55 years | 130 (14.4%) | 21 (25.9%) | 10 (25.6%) | 151 (15.3%) |
| 56 - 69 years | 44 (4.9%) | 14 (17.3%) | 6 (15.4%) | 58 (5.9%) |
| ≥ 70 years | 7 (0.8%) | 3 (3.7%) | 1 (2.6%) | 10 (1.0%) |
| **Reasons for Consultation** |  |  |  |  |
| Discuss results | 834 (92.4%) | 41 (50.6%) | 20 (51.3%) | 875 (88.9%) |
| Understand how to treat thyroid dysfunction | 497 (55.0%) | 29 (35.8%) | 19 (48.7%) | 526 (53.5%) |
| Interested in starting medication | 375 (41.5%) | 20 (24.7%) | 17 (43.6%) | 395 (40.1%) |
| Currently taking medication and want to discuss if changes are necessary | 38 (4.2%) | 45 (55.6%) | 13 (33.3%) | 83 (8.4%) |
| Not Sure | 30 (3.3%) | 1 (1.2%) | 0 (0.0%) | 31 (3.2%) |
| **Personal History** |  |  |  |  |
| Had Personal Thyroid History | 157 (17.4%) | 48 (59.3%) | 21 (53.8%) | 205 (20.8%) |
| Received thyroid surgery, | 17 (1.9%) | 7 (8.6%) | 3 (7.7%) | 24 (2.4%) |
| radioactive iodine therapy, or radiation of the head and neck |  |  |  |  |
| **Diagnosed with a chronic condition §** |  |  |  |  |
| Any chronic condition | 120 (13.3%) | 10 (12.3%) | 7 (17.9%) | 130 (13.2%) |
| **Family History ¶** |  |  |  |  |
| Hypothyroidism | 367 (40.6%) | 40 (49.4%) | 20 (51.3%) | 407 (41.4%) |
| Hyperthyroidism | 108 (12.0%) | 8 (9.9%) | 4 (10.3%) | 116 (11.8%) |
| Hashimoto's (autoimmune) thyroiditis | 106 (11.7%) | 12 (14.8%) | 6 (15.4%) | 118 (12.0%) |
| **Number of Symptoms ¶¶** |  |  |  |  |
| Mean number of symptoms (s.d.) | 6.09 (3.39) | 5.37 (3.49) | 5.00 (2.48) | 6.03 (3.41) |

Data are the number and percentage of those within a telemedicine category unless otherwise indicated. Individuals could endorse more than one reason for opting into telemedicine consultation
\* Non-treatment telehealth consultations were available to all Thyroid test takers
† Treatment telemedicine consultation eligibility: TSH value $\geq 4$ mIU/L or currently taking thyroid medications and had no contraindications
‡ Only patients receiving a treatment telemedicine consultation were eligible to be prescribed thyroid medication
§ Chronic conditions included anemia, celiac disease, type I diabetes, and "other"
¶ The three most common family history conditions were included in this table, other conditions included chronic immune thyroiditis, goiter, and Graves disease
¶¶ Index of the number of symptoms, associated with both hypo and hyperthyroidism, each patient reported. The most commonly reported symptoms were: fatigue and weakness, weight gain, intolerance to cold, depression, impaired memory, and nerve entrapment.

The number of reported chronic conditions was similar among non-treatment and treatment telehealth patients (Table 3). The most common chronic condition among all telehealth patients was anemia (n=70, 7.1%) followed by celiac disease (n=18, 1.8%) and type 1 diabetes (n=4, 0.4%). The most common family history condition was hypothyroidism, followed by Hashimoto’s thyroiditis (i.e., autoimmune hypothyroidism) and hyperthyroidism (Table 3).

The average number of symptoms reported among all telehealth users was 6 (s.d.=3, Table 3). The most common symptom reported was fatigue and weakness (n=768, 78%), which was most often co-reported with weight gain (n=621, 63.1%), intolerance to cold (n=444, 45.1%), depression (n=438, 44.5%), impaired memory (n=437, 44.4%).

## Discussion

This pilot program paired consumer initiated at-home sample collection and lab-based testing with the option for follow-up telehealth consultations. It was utilized by individuals interested in pursuing new thyroid diagnoses as well as by those with preexisting thyroid disorders who were likely partial to the ability to access care based on individual time and location preferences. In this pilot program, 16% of those whose lab values were indicative of overt or subclinical thyroid function engaged in telehealth consultations. Over half of those that opted into a telemedicine treatment consultation or were prescribed treatment had a personal history of thyroid problems. Additionally, over 50% were taking thyroid medication at the time of testing and wanted to discuss that with a clinician. These numbers highlight patient acceptance and interest in telehealth consultations as an adjunct to consumer initiated at-home thyroid tests and for ongoing treatment and monitoring of previously diagnosed thyroid disorders.

Studies are lacking on the use of telemedicine as a substitute for in-person visits with a provider for thyroid care. Measurement of TSH is an effective first step in assessment of thyroid function in most patients^14^ and although limitations exist, many aspects of the physical exam can be conducted via telehealth with the assistance of the patient such as visualization of the thyroid, eyes, skin, and assessment for tremor.^5^ Overall, paired with laboratory testing and referral to in-person follow-up procedures for conditions needing specialized attention, telemedicine is becoming a more acceptable option for thyroid care.^5, 6^ This may be particularly true for routine monitoring of thyroid conditions and appointments for prescription refills or dose adjustments where diagnostic testing beyond lab values is not necessarily indicated. This aligns with a large proportion of consumers’ reasons for requesting a telemedicine consultation in our sample.

Importantly, it is estimated that 30-40% of individuals reporting thyroid disease or taking thyroid medications continue to have abnormal TSH levels.^15-17^ Indeed, in our sample, 31% of those currently on thyroid medications fell into clinical thyroid categories. This emphasizes the importance of continued monitoring in patients taking thyroid medications and for compliance or concerns of misuse. Additionally, repeated lab measurements are recommended for thyroid dysfunction diagnosis, particularly of subclinical disease.^14, 18-21^ However, patients often experience prolonged wait times for appointments to see medical providers.^22^ The adult endocrinology subspecialty has a particularly large burden of mismatch between provider supply and patient demand that is forecasted to widen.^23, 24^ Remote testing models that pair at-home sample collection for certified laboratory testing with telehealth consultations can help to limit the bottleneck for in-person thyroid care providing a convenient, accurate, efficient, and rapid option for diagnosis, routine monitoring, and medication adjustments.

The popularity of CIT offerings is evident with over 8,000 customers taking part in this 10-week pilot with just under 1,000 receiving in telehealth consultations. We observed on average 16% telehealth engagement in individuals falling into one of the four the thyroid dysfunction categories, which may reflect interest yet ongoing hesitancy among patients regarding engagement in telemedicine. In a nationwide survey conducted in 2015, 52% of respondents replied they would be willing to see their own healthcare provider via telemedicine while just under 20% were willing to see a different provider from a different organization via telemedicine.^25^ This aligns with our program, which involved consultation with a new provider. In addition to preferring to see their own provider, barriers to telemedicine adoption in this model of care include limitations in technology, fear of the use of technology and data security, and wanting the provider to have access to their health records.^25, 26^

Compared to estimates for the U.S. population,^16^ this sample was enriched for cases of overt hypothyroidism (0.9% this sample vs 0.3% NHANES) and subclinical hyperthyroidism (3% vs 0.7%). Individuals choosing to utilize the CIT/telehealth program were likely self-selecting given pre-existing concerns of thyroid dysfunction. Indeed, those that opted into the telehealth consultations commonly reported family history of thyroid disorders, had an average of six thyroid related symptoms, and 40% indicated interest in starting a thyroid medication. Thus, while not representative of a truly random population sample, the sample is adequate size to assess and describe major features among those interested in utilizing CIT to manage thyroid-related health concerns. Overall, there is significant potential for a CIT/telehealth program to provide increased access to care for both early detection and continued monitoring of thyroid dysfunction and facilitate active patient participation in health care decision making and self-management.

### Limitations

This analysis of CIT data has limitations. Lack of race or ethnicity information and limited representation of males precluded meaningful sub-analyses. In addition, thyroid function was based on single, instead of repeat, diagnostic measurement. Multi-laboratory testing also poses variance in independently determined reference ranges which complicates quantitative analyte analysis and thresholds for thyroid function categorization or overt diagnosis were lab specific. Further, there remains contention within the field regarding the appropriate thresholds for normal TSH values, particularly within subgroups such as older adults or specific racial and ethnic groups.^27-30^ We utilized thresholds based on traditional thyroid dysfunction categories and each laboratory’s unique reference ranges. For comparison, the United States Preventive

Services Task Force (USPSTF) suggests screening and initiation of treatment for TSH values > 10 mlU/L for hypothyroidism and undetectable or < 0.1 mlU/L for overt hyperthyroidism.^18^ Of note, the average TSH result among patients who received a prescription medication through this program was 9.66 mIU/L (s.d.=13.20), just below the USPSTF recommendations. Unfortunately, it was unknown whether the prescriptions written as part of this program were dose adjustments, refills, or for a new medication. Finally, personal objectives for testing or rate of in-person physician follow-up for those that did not engage in the offered telehealth program were not available. This information, particularly for those falling within clinical categorization for potential treatment, would have been useful in understanding the patient motivations, the barriers to their adoption of telehealth, and in designing new features aimed at improving telemedicine engagement in those needing follow-up care based on their lab results.

## Conclusions

These findings demonstrate that expanding options for screening and obtaining ongoing care for monitoring of chronic conditions through consumer-initiated telemedicine programs is an innovative new model that is prime for development. Models that pair at-home lab testing with telemedicine consultations within the same provider group for continuity of treatment records are well-positioned to be the leader in this new healthcare delivery space. We conclude that adjoining CIT, at-home sample collection, and telehealth is a potentially useful and innovative approach for expanding access to early detection of thyroid dysfunction in those experiencing symptoms and monitoring thyroid health in those with an established diagnosis or prescription. Increasing utilization of post-test telemedicine consultation, particularly for individuals with values indicative of overt thyroid dysfunction, is a critical next step.

## Data Availability

All data produced in the present study are available upon reasonable request to the authors

## Acknowledgements

We would like to acknowledge Dr. Natalie Daumeyer, Dr. Doug Elwood,, and Devon Humphreys for their assistance with preparing and providing feedback on the manuscript. This work analysis was funded by Everly Health, Inc. Portions of the data included in this manuscript were presented at the 90th Annual Meeting of the American Thyroid Association, September 30 to October 3, 2021.

## Funding Acknowledgement

This research received no specific grant from any funding agency in the public, commercial, or not-for-profit sectors, but was sponsored by Everly Health, Inc.

